# Genomic Epidemiologic Investigation of a Multispecies Hospital Outbreak of NDM-5-Producing Enterobacterales Infections

**DOI:** 10.1101/2023.08.31.23294545

**Authors:** Nathan J. Raabe, Abby L. Valek, Marissa P. Griffith, Emma Mills, Kady Waggle, Vatsala Rangachar Srinivasa, Ashley M. Ayres, Claire Bradford, Hannah Creager, Lora L. Pless, Alexander J. Sundermann, Daria Van Tyne, Graham M. Snyder, Lee H. Harrison

**Affiliations:** Microbial Genomic Epidemiology Laboratory, Center for Genomic Epidemiology, University of Pittsburgh, 3507 Victoria Street, BST-10 E1000-4A, Pittsburgh, Pennsylvania 15213, USA; Division of Infectious Diseases, University of Pittsburgh School of Medicine, 3550 Terrace Street, 818 Scaife Hall, Pittsburgh, Pennsylvania 15261, USA; Department of Epidemiology, School of Public Health, University of Pittsburgh, 130 De Soto Street, Pittsburgh, Pennsylvania 15261, USA; Department of Infection Control and Hospital Epidemiology, UPMC Presbyterian, 200 Lothrop Street, Pittsburgh, Pennsylvania 15213, USA; Department of Pathology, University of Pittsburgh Medical Center, 200 Lothrop Street Pittsburgh, PA 15213; Department of Pathology, University of Pittsburgh School of Medicine, 200 Lothrop St, S-417 BST, Pittsburgh, PA 15261

**Keywords:** New-Delhi metallo-β-lactamase, Outbreak Investigation, Plasmid, Whole genome sequencing

## Abstract

**Background:** New Delhi metallo-β-lactamase (NDM) represents an emergent mechanism of carbapenem resistance associated with high mortality and limited antimicrobial treatment options. Because the *bla*_NDM_ resistance gene is often carried on plasmids, traditional infection prevention and control (IP&C) surveillance methods like speciation, antimicrobial resistance testing, and reactive whole genome sequencing (WGS) may not detect plasmid transfer in multispecies outbreaks.

**Methods:** Initial outbreak detection of NDM-producing Enterobacterales identified at an acute care hospital occurred via traditional IP&C methods and was supplemented by real-time WGS surveillance, which was performed weekly using the Illumina platform. To resolve NDM-encoding plasmids, we performed long-read Oxford Nanopore sequencing and constructed hybrid assemblies using Illumina and Nanopore sequencing data. Reports of relatedness between NDM-producing organisms and reactive WGS for suspected outbreaks were shared with the IP&C team for assessment and intervention.

**Findings:** We observed a multispecies outbreak of NDM-5-producing Enterobacterales isolated from 15 patients between February 2021 and February 2023. The 19 clinical and surveillance isolates sequenced included seven bacterial species and each encoded the same NDM-5 plasmid, which showed high homology to NDM plasmids previously observed in Asia. WGS surveillance and epidemiologic investigation characterized ten horizontal plasmid transfer events and six bacterial transmission events between patients housed in varying hospital units. Transmission prevention focused on enhanced observation and adherence to basic infection prevention measures.

**Interpretation:** Our investigation revealed a complex, multispecies outbreak of NDM that involved multiple plasmid transfer and bacterial transmission events, increasing the complexity of outbreak identification and transmission prevention. Our investigation highlights the utility of combining traditional IP&C and prospective genomic methods in identifying and containing plasmid-associated outbreaks.

**Funding:** This work was funded in part by the National Institute of Allergy and Infectious Diseases, National Institutes of Health (NIH) (R01AI127472) (R21AI1783691).

**Summary:** We investigated a multispecies outbreak of Enterobacterales harboring the same New-Delhi metallo-β-lactamase-encoding plasmid using both traditional infection prevention and genomic approaches. Our investigation revealed a complex outbreak involving 7 bacterial species, including both bacterial transmission and plasmid transfer events.

## INTRODUCTION

Bacteria that cause healthcare-associated infections (HAIs) can harbor plasmids encoding genes conferring antibiotic resistance to commonly prescribed antimicrobial agents, leading to pathogens that are well adapted to the hospital environment. Infections with antimicrobial-resistant pathogens are associated with poor patient outcomes and have limited options for therapy.^1^ Horizontal plasmid transfer represents the primary mechanism by which antibiotic resistance genes can be shared between different bacterial species.^2^ Horizontal gene transfer (HGT) of plasmids containing resistance genes can occur within a co-colonized patient or a hospital environment contaminated with different bacterial species, and can result in transmission of different bacterial species harboring the same resistance plasmid to other hospitalized patients.

New Delhi metallo-β-lactamase (NDM), encoded by *bla*_NDM_, is an emerging mechanism of carbapenem resistance that is seen in HAIs. First isolated in 2008, NDM is associated with an increased risk of mortality and limited therapeutic options.^3^ Numerous *bla*_NDM_ variants have been found to be encoded by a variety of plasmid types, and dozens of different NDM variants have been identified in more than 60 host species spanning multiple bacterial families.^4^

Whole genome sequencing (WGS) is frequently used to identify and investigate hospital-associated bacterial outbreaks.^5, 6^ Existing methods largely focus on genome-wide comparisons between isolates to assess the likelihood of transmission for the same bacterial species. However, WGS as used in outbreak investigations is generally not designed to identify plasmid transfer among different bacterial species. Therefore, detecting multi-species plasmid-associated outbreaks presents a considerable challenge that requires tailored approaches to identify both plasmid transfer and bacterial transmission.

Globally and within the United States, the incidence of NDM-producing infections per year is increasing.^7^ Prior studies of NDM outbreaks have involved both single and multiple bacterial species.^8–10^ In this study, we describe the features of a multi-species, plasmid-associated NDM outbreak at our institution that was detected and investigated by both epidemiologic and genomic methods.

## RESEARCH IN CONTEXT

### Evidence before this study

We conducted a literature search using the PubMed database for genomic epidemiology studies utilizing whole genome sequencing (WGS) to investigate plasmid-associated outbreaks of carbapenemase producing Enterobacterales from database inception to July 21, 2023. The search terms used were ((NDM) OR (New-Delhi metallo-β-lactamase) OR (carbapenem-resistant) OR (carbapenemase)) AND (Enterobacterales) AND (infection control) AND (transmission) AND (healthcare OR hospital) AND (outbreak) AND (plasmid) AND ((WGS) OR (whole genome sequencing)). Prior studies have employed WGS to retrospectively investigate both single and multi-species outbreaks of Enterobacterales harboring plasmids with carbapenemase genes. In tandem with epidemiological data, WGS has recently shown promise in providing evidence of both bacterial transmission and plasmid transfer in plasmid-associated outbreak investigations. However, all studies to date have analyzed such outbreaks retrospectively and are thus likely unable to capture the real-time dynamics of an ongoing outbreak. This may result in delayed detection, response, and implementation of infection control measures, thus missing opportunities for timely and targeted intervention. As a result, there exists a gap in the literature regarding the need for real-time strategies to effectively identify and contain plasmid-associated outbreaks.

### Added value of this study

This study builds on the retrospective approach by implementing real-time WGS surveillance, allowing for early detection of plasmid-associated outbreaks and tracking plasmid transfer events between different bacterial species. To our knowledge, this is the first study utilizing a real-time approach which combines WGS surveillance and epidemiological data to identify both bacterial transmission and horizontal plasmid transfer in a multispecies outbreak of Enterobacterales harboring the same NDM-encoding plasmid. A real-time collaborative approach allowed for timely and targeted interventions to halt the spread of the outbreak plasmid. This study provides evidence which supports the use of real-time WGS surveillance, epidemiological data, and targeted interventions to describe and contain plasmid-associated outbreaks of drug-resistant bacteria.

### Implications of all the available evidence

The detection and control of dangerous antibiotic resistance plasmids requires a nuanced approach which captures inter-species plasmid transfer in addition to bacterial transmission events. The spread of important resistance-encoding plasmids goes undetected by current outbreak investigation methods which only use WGS to detect clonal transmission. To date, studies investigating multispecies plasmid-associated outbreaks have only identified interspecies transfer during retrospective analysis of WGS combined with epidemiological data. Our study suggests the utility of approaches which use real-time WGS surveillance alongside epidemiological data to both describe and halt the spread of antibiotic resistance plasmids using targeted interventions in a hospital setting.

## METHODS

### Study Setting

This study was performed at UPMC Presbyterian Hospital, an adult tertiary care facility with 758 beds, including 134 critical care beds, and where over 400 solid organ transplants are performed annually. In November 2021, we instituted real-time WGS surveillance for key bacterial pathogens as part of the Enhanced Detection System for Healthcare-Associated Transmission (EDS-HAT), which was designed to identify hospital outbreaks missed by traditional infection prevention and control (IP&C) methods.^5, 11^ EDS-HAT uses a combination of WGS surveillance and machine learning of the electronic health record to detect outbreaks and identify the most likely transmission route, respectively. Ethics approval was obtained from the University of Pittsburgh Institutional Review Board (STUDY21040126).

### Study Isolates

Study isolates were from patient cultures obtained from February 2021 through February 2023 for either clinical care purposes or IP&C active surveillance of patient colonization as part of the outbreak investigation. Select high-priority healthcare-associated bacterial pathogens were collected for WGS surveillance as part of EDS-HAT.^12^ Inclusion criteria for isolates identified for WGS surveillance were: i) patient hospital admission date was greater than two days before the culture date, or ii) the patient had an inpatient or outpatient encounter at a UPMC facility in the 30 days before the culture date.

### Microbiologic Methods

Isolates were sub-cultured onto a blood agar plate for species identification and antimicrobial susceptibility testing by the clinical microbiology laboratory. The clinical microbiology laboratory routinely tests carbapenem-resistant Enterobacterales for carbapenemase production using the modified carbapenem inactivation test, which is performed as described in the Clinical Laboratory Standard’s Institute’s Performance Standards for Antimicrobial Susceptibility Testing (M100) document^13^ except that a 10 µL loop was used to prepare the inoculum. Because this test requires overnight incubation, isolates testing non-susceptible to ceftazidime/avibactam and/or meropenem/vaborbactam are flagged by the automated antimicrobial susceptibility test system software and brought to the attention of the medical director. Early in the outbreak, the presence of NDM in these isolates was confirmed by PCR for *bla*_NDM_ performed in a research laboratory; subsequently, this method was replaced by confirmation testing performed in the clinical microbiology laboratory using the NG-Test CARBA 5 lateral flow assay (Hardy Diagnostics, Santa Maria CA).

### Whole Genome and Plasmid Sequencing

Isolates were streaked onto a blood agar plate (BD, Franklin Lakes, NJ) and grown overnight at 37°C in preparation for DNA extraction by the Microbial Genomic Epidemiology Laboratory. DNA was extracted from overnight cultures of a single bacterial colony using the MagMax DNA Multi-Sample Ultra 2.0 extraction kit on a KingFisher Apex (Thermo Fisher Scientific). WGS was performed using the Illumina NextSeq 550 platform (Illumina, San Diego CA), with libraries prepared using an Illumina DNA Prep Tagmentation Kit on an EpMotion (Illumina, San Diego CA) and sequenced using 2 ×150 bp paired-end reads. Bacterial species was determined by genomic comparisons with typed strains of suspected species using fastANI.^14^ Reads were assembled using SPAdes v3.15.5^15^ and genomes were annotated with Prokka v1.14.5.^16^ WGS data quality control was preformed using QUAST v5.2.0 and if any sample had less than 35x coverage, it was not considered for further analyses.^17^ Multi-locus sequence type (ST) was determined using PubMLST (https://github.com/tseemann/mlst). Core genome pairwise single nucleotide polymorphisms (SNP) differences were determined using Snippy v4.6.0 (https://github.com/tseemann/snippy).

For optimal characterization of plasmids, we performed long-read sequencing using a MinION device with R9.4.1 flow cells (Oxford Nanopore Technologies, Oxford, United Kingdom). Genomic DNA prep for long-read sequencing was obtained either from the same gDNA prep used for Illumina sequencing or was extracted again with a DNeasy Blood and Tissue Kit (Qiagen, Hilden, Germany). Libraries were constructed using a rapid multiplex barcoding kit (catalog number SQK-RBK004). Base-calling and read processing were performed using Guppy v6.3.9 (Oxford Nanopore Technologies, Oxford, United Kingdom) using default parameters. Hybrid assembly based on both Illumina and MinION sequencing data was performed using Unicycler v0.5.0.^18^ Plasmids were identified as closed, circular contigs encoding identifiable plasmid replicons using BLASTn and the PlasmidFinder database.^19^ For comparison of outbreak *bla*_NDM-5_-harboring plasmids, publicly available sequencing data for *IncX3* family plasmids harboring *bla*_NDM-5_ were obtained from NCBI and included in this study based on a similarity score, determined by multiplying the percent coverage and percent identity of each plasmid. Forty-seven plasmids with similarity scores >99·9% were included for comparison. Comparative genomic analysis on plasmid sequences was performed by Roary v3.13.0^20^ based on Prokka v1.14.5 annotations.^16^ A phylogenetic tree of plasmid sequences was constructed using RAxML-HPC v8.2.12^21^ with 100 bootstraps from the core genome alignment produced by Roary.^20^ Plasmid annotations were created using Bakta (https://github.com/oschwengers/bakta) and visualized in Geneious 2023.1.2 (http://www.geneious.com/). Hybrid genome assemblies generated in this study were submitted to NCBI under BioProject PRJNA981541.

### Definitions of the Outbreak Plasmid, Horizontal Plasmid Transfer, and Bacterial Transmission

The outbreak plasmid was defined as an *IncX3* family conjugative plasmid harboring *bla*_NDM-5_ that had sequence coverage values (i.e., the proportion of each plasmid’s sequence that was found in the Illumina genome) of at least 95·0%, and sequence identity of <15 SNPs per 100kb of plasmid sequence compared with the fully resolved, circular plasmid sequence recovered from an early outbreak isolate.^22^ An *intra-patient horizontal plasmid transfer event* was defined as a patient from whom two different bacterial species containing the outbreak plasmid were isolated, either on the same or different days. An *inter-patient horizontal plasmid transfer event* was defined as two patients from whom different bacterial species, or same species but different genetic lineages, carrying the outbreak plasmid were isolated, supported by epidemiologic evidence of transfer. A *bacterial transmission event* was defined as two or more patients whose isolates were of the same bacterial species and differed by a small number of core genome SNPs. Based on our own experience and the literature, we used an initial SNP cut-off of ≤15 to determine bacterial transmission but allowed for up to 19 SNPs if supported by epidemiologic evidence.^5, 23–33^

### Epidemiologic Investigation and Intervention

The IP&C team investigated prior hospital locations, surgeries or other procedures, shared medical equipment, and staff shared among patients with carriage (infection or colonization) of the outbreak plasmid by reviewing the electronic medical record and detailed investigation for potential associations. Active surveillance among potentially exposed patients was performed by obtaining a peri-anal swab and identifying NDM-carrying Enterobacterales using the microbiologic methods described above.

Patients were considered potentially exposed if the patient was on the same inpatient unit for at least one concomitant day as an index patient with NDM carriage (carriage was considered to begin with the date of the first positive clinical or active surveillance culture) or within three days of the index patient’s transfer from the unit. Active surveillance among potentially exposed patients was performed approximately weekly, for at least one round during or after exposure, and repeated at the discretion of IP&C dependent upon the index patient’s length of stay, clinical condition (relative to transmission risk), and prior active surveillance results. In the case of three index patients, only one round of active surveillance among potentially exposed patients was performed, and in the case of two index patients, active surveillance was not performed among potentially exposed patients as the index patients’ NDM isolates were identified approximately one year prior to identification of the outbreak, with no subsequent index patient encounters in that interval or during the investigation.

Interventions to prevent transmission focused on enhanced adherence to basic infection prevention measures. The IP&C team performed staff education related to NDM carriage and transmission, notification of investigation findings with leadership and front-line staff to increase engagement, audits with immediate and periodic feedback of adherence to hand hygiene and personal protective equipment use, limiting reusable medical equipment, and enhanced environmental cleaning defined as supervised terminal room cleaning and increased frequency of routine cleaning of index patient inpatient room. An observer was assigned to each index patient after NDM infection or carriage was identified; the observer documented each healthcare worker entering the inpatient room each shift and enforced hand hygiene, personal protective equipment use, and reusable medical equipment-related practices. The observer was assigned to travel when feasible with the patient for off-unit clinical care. Interventions were initiated immediately with each identification and notification of a new NDM-positive patient.

## RESULTS

### Description of the Outbreak

An outbreak investigation using traditional IP&C methods began in April 2022 when it was noted that four NDM-producing carbapenem-resistant organisms belonging to different species (*Raoultella planticola*, NDM-1; *Proteus mirabilis* and *Morganella morganii*, NDM-5; *Klebsiella aerogenes,* NDM-5) were isolated from three patients between March and April. Analysis of WGS data concluded that two isolates (*P. mirabilis* and *M. morganii*) collected from one patient harbored a *bla*_NDM-5_ encoding IncX family plasmid. Additional collection and WGS of carbapenemase-producing isolates over the following nine months identified a multispecies outbreak of NDM-5-producing Enterobacterales isolated from 15 patients between February 2021 and February 2023, all of which contained the same outbreak plasmid (Figure 1, Table 1). Four patients with NDM-producing isolates detected during the same period were found to harbor isolates with non-outbreak NDM-1-carrying plasmids and were excluded from this investigation.

**Figure 1.**
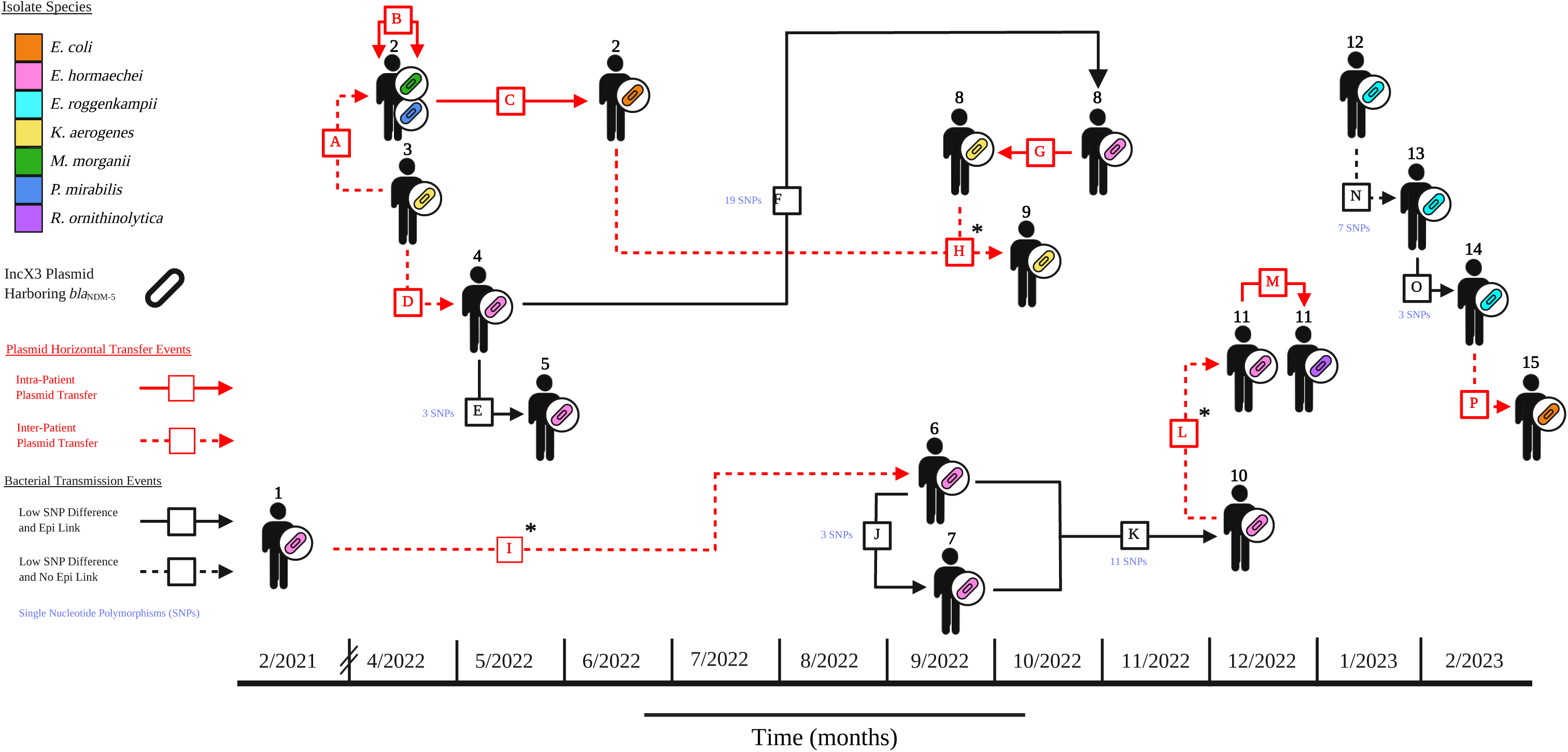
Plasmid transfer and bacterial transmission events among patients with bacterial isolates found to harbor the outbreak plasmid. Patients 1-15 are shown in chronological order based on date of first positive culture for an isolate found to harbor the outbreak plasmid. Bacterial species are represented by colors on the key. Intra- and inter-patient horizontal plasmid transfer events are shown by solid and dashed red arrows, respectively (see text for definitions). Bacterial transmission events are shown as dashed (indicating no known epidemiological link) or solid black arrows (an IP-confirmed epidemiological link). Presumed directionality of the events are indicated by the arrowheads, with double-sided arrows referring to unknown directionality. Pairwise differences in core genome single nucleotide polymorphisms (SNPs) were utilized to measure the genetic relatedness of isolates. Low SNP differences are denoted in blue alongside bacterial transmission arrows. *Note:* *, Indicates horizontal plasmid transfer from one lineage of a species to another lineage of the same species.

**Table 1.** NDM-5-encoding outbreak isolates.

| **Isolate No.** | **NCBI Accession** | **Patient No.** | **Unit First Positive** | **Culture Month** | **Species** | **Source** |
| --- | --- | --- | --- | --- | --- | --- |
| 1 | SAMN35677778 | 1 | Med/Surg A | 2/2021 | *Enterobacter hormaechei* | Wound |
| 2 | SAMN35677779 | 2 | Med/Surg B | 4/2022 | *Proteus mirabilis* | Bone |
| 3 | SAMN35677780 | 2 | Med/Surg B | 4/2022 | *Morganella morganii* | Bone |
| 4 | SAMN35677781 | 2 | Med/Surg B | 6/2022 | *Escherichia coli* | Peri-anal Swab |
| 5 | SAMN35677782 | 3 | Med/Surg C | 4/2022 | *Klebsiella aerogenes* | Urine |
| 6 | SAMN35677783 | 4 | Med/Surg D | 5/2022 | *Enterobacter hormaechei* | Blood |
| 7 | SAMN35677784 | 5 | ICU A | 5/2022 | *Enterobacter hormaechei* | Blood |
| 8 | SAMN35677785 | 6 | OP Surg | 9/2022 | *Enterobacter hormaechei* | Tissue |
| 9 | SAMN35677786 | 7 | Med/Surg E | 9/2022 | *Enterobacter hormaechei* | Peri-anal Swab* |
| 10 | SAMN35677787 | 8 | ICU B | 9/2022 | *Klebsiella aerogenes* | Peri-anal Swab† |
| 11 | SAMN35677788 | 8 | IP Rehab | 11/2022 | *Enterobacter hormaechei* | Urine† |
| 12 | SAMN35677789 | 9 | ICU B | 10/2022 | *Klebsiella aerogenes* | Peri-anal Swab† |
| 13 | SAMN35677790 | 10 | ICU B | 12/2022 | *Enterobacter hormaechei* | Peri-anal Swab† |
| 14 | SAMN35677791 | 11 | Med/Surg F | 12/2022 | *Enterobacter hormaechei* | Wound |
| 15 | SAMN35677792 | 11 | Med/Surg F | 12/2022 | *Raoultella ornithinolytica* | Wound |
| 16 | SAMN35677793 | 12 | Med/Surg G | 1/2023 | *Enterobacter roggenkampii* | Sputum |
| 17 | SAMN35677794 | 13 | Med/Surg H | 2/2023 | *Enterobacter roggenkampii* | Blood |
| 18 | SAMN35677795 | 14 | IP Rehab | 2/2023 | *Enterobacter roggenkampii* | Peri-anal Swab‡ |
| 19 | SAMN35677796 | 15 | ICU C | 2/2023 | *Escherichia coli* | Peri-anal Swab‡ |

NCBI, National Center for Biotechnology Information; Med/Surg, Medical/Surgical Unit; ICU, Intensive Care Unit; OP Surg, outpatient surgery; IP Rehab, Inpatient rehabilitation unit; NA, not applicable.
\*, identified by active surveillance for a non-outbreak NDM-1 enzyme
†, identified by routine active surveillance for carbapenem-resistant Enterobacteriaceae carriage.
‡, identified by active surveillance for the outbreak NDM-5 plasmid

Fifty-three rounds of active surveillance were performed around 13 index patients, ranging from one to 17 rounds (median, three rounds). Of 1,198 individuals meeting criteria for active surveillance, 913 (76·2%) had a peri-anal swab obtained. Among these, 2 (0·2%) were positive for the outbreak NDM-5-encoding plasmid; one additional case of outbreak NDM-5 plasmid carriage was identified during active surveillance around patients with non-outbreak NDM-1 enzyme carriage. Three additional patients were identified while performing routine active surveillance for carbapenem-resistant organisms (Table 1).

Outbreak isolates were obtained from patients in a variety of acute care units at the time of clinical or active surveillance culture (Table 1). Positive cultures were most commonly obtained from peri-anal surveillance swabs (n=7), and the 19 outbreak isolates comprised seven bacterial species, most commonly *Enterobacter hormaechei* (n=8), each encoding the same NDM-5 plasmid (Table 1). Three (15·8%) patients died within 30 days of their positive culture, however only one potentially attributable to infection with an outbreak pathogen. As of August 2023, no additional isolates harboring the NDM-5 plasmid have been identified.

### Plasmid Identification and Analysis

Hybrid genome assemblies of isolates in the outbreak revealed that *bla*_NDM-5_ was carried on a 46·2 kb *IncX3* family conjugative plasmid (Supplemental Figure 1). The plasmid sequence was highly conserved between isolates, with 0-2 SNPs identified between plasmids among outbreak isolates. Two plasmids differing in length, pEB00162 (55·781 kb) (Isolate 13) and pENTC0964 (45·198 kb) (Isolate 1), showed acquisition or loss of a transposase gene, respectively. Among the sequence data obtained from NCBI for 47 isolates with >99·9% similarity to the outbreak plasmid, 39 (83·0%) were from China, 37 (78·7%) were *Escherichia coli*, and 33 (70·2%) were isolated from humans (Figure 2, Supplemental Table 1). The outbreak plasmid described here clustered most closely with plasmids isolated from China between 2013 and 2019.

**Figure 2.**
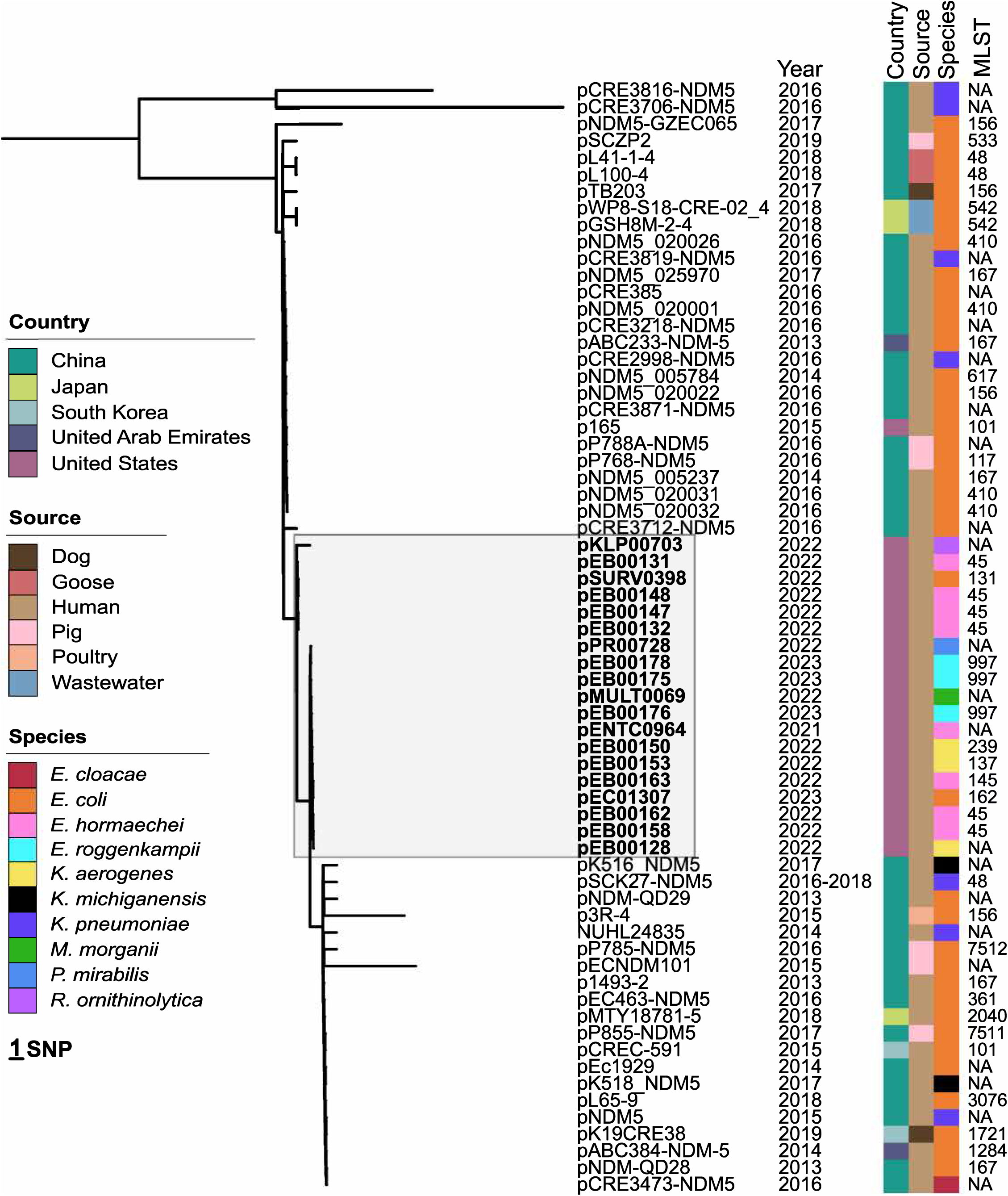
UPMC outbreak plasmids in the global context. *IncX3 bla*NDM-5-encoding plasmid sequences from 19 UPMC isolates (bolded and shaded in grey) were compared to 47 plasmid sequences gathered from NCBI with a similarity score (% identity multiplied by % coverage) of ≥ 99·9%. The midpoint rooted tree was constructed using RAxML HPC with 100 bootstraps based on a core genome alignment, consisting of 61 core genes, produced by Roary. Undefined or unavailable MLSTs are noted as not available (NA).

### Horizontal Plasmid Transfer and Transmission Events

WGS analysis combined with IP&C investigation identified 16 total plasmid transfer or bacterial transmission events across various hospital units (Figure 1). These 16 events included six bacterial transmissions, six inter-patient plasmid transfers, and four intra-patient plasmid transfers (Table 2). Suspected transmission routes were identified for 5/6 (83·3%) bacterial transmissions. Plasmid transfer events required an epidemiological linkage between patients by definition; we identified a single possible source and route for 9/10 (90·0%) plasmid transfer events. For Patients 2 (Events B and C), 8 (Event G), and 11 (Event M), who tested positive for multiple isolates containing the outbreak plasmid, intra-patient horizontal plasmid transfers were inferred to have occurred during co-colonization with two different bacterial species. We were unable to identify the source patient for two patients (Patient 3 and Patient 12) excluding the outbreak index patient and could not identify a transmission route for 1 event (Event N). IP&C interventions for events with known transmission routes are summarized in Table 2.

**Table 2.** Details of transmission events.

| Event | Event Date | Event Type | Putative Source / Recipient Patient ID | Patient ID<br>Bacterial Species | Core Bacteria<br>Genome SNPs | Hypothesized Bacterial Transmission / Plasmid Transfer Route | Route-specific IP&C Intervention |
| --- | --- | --- | --- | --- | --- | --- | --- |
| **A** | Mar. – Apr. 2022 | Inter-patient plasmid transfer | 3 → 2 | 3) *K. aerogenes*<br>2) *M. morganii* or *P. mirabilis* | NA | Shared unit, shared ancillary staff (within 1 hour) | Education to unit reinforcing basic infection prevention practices. |
| **B** | Apr. – Jun. 2022 | Intra-patient plasmid transfer | 2 ↔ 2 | 2) *P. mirabilis*<br>2) *M. morganii* | NA | Co-colonization | NA |
| **C** | Apr. – Jun. 2022 | Intra-patient plasmid transfer | 2 → 2 | 2) *P. mirabilis* or *M. morganii*<br>2) *E. coli* | NA | Co-colonization | NA |
| **D** | Apr. – May. 2022 | Inter-patient plasmid transfer | 3 → 4 | 3) *K. aerogenes*<br>4) *E. hormaechei* | NA | Shared specialized nursing service | Targeted education reinforcing basic infection prevention practices. |
| **E** | May. 2022 | Bacterial Transmission | 4 → 5 | 4) *E. hormaechei*<br>5) *E. hormaechei* | 3 | Shared specialized nursing service (within 1 hour) | Targeted education reinforcing basic infection prevention practices. |
| **F** | Sept. 2022 | Bacterial Transmission | 4 → 8 | 4) *E. hormaechei*<br>8) *E. hormaechei* | 19 | Shared specialized nursing service. | Targeted education reinforcing basic infection prevention practices. |
| **G** | Sept. – Nov. 2022 | Intra-patient plasmid transfer | 8 → 8 | 8) *K. aerogenes*<br>8) *E. hormaechei* | NA | Co-colonization | NA |
| **H** | Oct. 2022 | Inter-patient plasmid transfer | 2 → 9 | 2) *E. coli*<br>9) *K. aerogenes* | NA | Shared specialized nursing service. | Targeted education reinforcing basic infection prevention practices. |
|  |  |  | 8 → 9 | 8) *K. aerogenes*<br>9) *K. aerogenes* | 123,415 | Shared ancillary provider. |  |
| **I** | Jul. 2022 | Inter-patient plasmid transfer | 1 → 6 | 1) *E. hormaechei*<br>6) *E. hormaechei* | 25,404 | Shared procedural area | Targeted education reinforcing basic infection prevention practices. |
| **J** | Aug. 2022 | Bacterial Transmission | 6 → 7 | 6) *E. hormaechei*<br>7) *E. hormaechei* | 3 | Shared hospital unit (same time) | Supervised terminal clean of patient room by environmental services leader. Education to unit reinforcing basic infection prevention practices. |
| **K** | Sept. – Oct. 2022 | Bacterial Transmission | 6 → 10 | 6) *E. hormaechei*<br>10) *E. hormaechei* | 11 | Shared room in intensive care unit (different times) | Supervised terminal clean of patient room by environmental services leader. Education to unit reinforcing basic infection prevention practices. |
| **L** | Sept. 2022 | Inter-patient plasmid transfer | 10 → 11 | 10) *E. hormaechei*<br>11) *E. hormaechei* | 141,963 | Shared hospital unit (same time) | Supervised terminal clean of patient room by environmental services leader. Education to unit reinforcing basic infection prevention practices. |
| **M** | Dec. 2022 | Intra-patient plasmid transfer | 11 → 11 | 11) *E. hormaechei*<br>11) *R. ornithinolytica* | NA | Co-colonization | NA |
| **N** | Jan. – Feb. 2023 | Bacterial Transmission | 12 → 13 | 12) *E. roggenkampii*<br>13) *E. roggenkampii* | 7 | none identified | NA |
| **O** | Jan. – Feb. 2023 | Bacterial Transmission | 13 → 14 | 13) *E. roggenkampii*<br>14) *E. roggenkampii* | 3 | Shared hospital unit (same time) | Supervised terminal clean of patient room by environmental services leader. Education to unit reinforcing basic infection prevention practices. |
| **P** | Feb. 2023 | Inter-patient plasmid transfer | 14 → 15 | 14) *E. roggenkampii*<br>15) *E. coli* | NA | Shared room (different times) | Supervised terminal clean of patient room by environmental services leader. Education to unit reinforcing basic infection prevention practices. |

Horizontal plasmid transfer and bacterial transmission events A-P are designated as inter- or intra-patient plasmid transfer or bacterial transmission and listed alongside event date, event type, putative source and recipient patient, bacterial species, SNP difference, transmission/transfer route, and IP&C intervention. SNPs, refers to core genome pairwise single nucleotide polymorphisms (SNPs) differences between bacterial isolates; → and ↔, represent putative and unknown directionality of transfer/transmission event, respectively; NA, not applicable.

Not all isolates of the same species implicated in the outbreak were genetically related. Core genome SNP comparisons of same-species isolates ranged from 3 to 141,963 SNPs (Table 2). We observed three genetically related clusters – two *E. hormaechei* clusters (both n=3) and one *Enterobacter roggenkampii* cluster (n=3). Thus, bacterial transmission of genetically related isolates accounted for only 6/19 (31·6%) total events. Conversely, 13/19 (68·4%) of events involved either inter- or intra-patient plasmid transfer, and we observed three instances of inter-patient horizontal plasmid transfer occurring between genetically unrelated isolates of the same species, denoted by an asterisk (Figure 1).

## DISCUSSION

In this investigation, we characterized a two-year outbreak of an NDM-5 plasmid using WGS and traditional IP&C methods. We identified ten plasmid transfer events and six bacterial transmission events resulting in colonization or infection among 15 patients. Epidemiological linkages, when paired with genomic evidence, provided clear pathways for the transfer of the NDM-carrying plasmid between organisms and subsequent transmissions of plasmid-harboring bacteria between patients. Outside of within-patient transfers between different species, some plasmid transfers were unit-based transmission events; however, a substantial proportion may have been facilitated by care in multiple different units, in a non-unit shared location, or via transmission routes not identified. Our investigation demonstrates the challenge of identifying and preventing dissemination of antimicrobial resistance elements in the healthcare setting when they occur across bacterial species and in complex transmission pathways.

The integration of real-time WGS surveillance facilitated highly sensitive, rapid identification of outbreak isolates, complementing traditional IP&C methods. By aligning genomic and epidemiological data, the spread of the NDM-5 plasmid could be tracked through both horizontal transfer and bacterial transmission events. This collaborative approach has the potential to identify undetected transmission, and therefore improve the adherence and effectiveness of infection prevention interventions to prevent further spread. Thus, the implementation of real-time WGS-based surveillance alongside IP&C interventions is a useful strategy for containing the spread of plasmid-associated antimicrobial-resistant outbreaks.

We observed more plasmid transfer events than bacterial transmission events overall. However, when considering only inter-patient events, we observed equal numbers of inter-patient plasmid transfers and bacterial transmissions. Inter-patient plasmid transfer events were described with inherently less certainty than intra-patient plasmid transfer events, as inter-patient transfer requires the assumption that transfer occurred via the identified epidemiological linkage between the isolates of two separate patients, rather than within the same patient. Nonetheless, this investigation suggests that plasmid associated transmission events may be common, yet not well characterized by methods that only detect same-species transmission of the same strain or by current IP&C methods that do not routinely utilize genomic data.

A defining characteristic of this outbreak is the extent to which transmission did not occur in a single unit, which is uncommon for descriptions of outbreaks due to carbapenem-resistant pathogens.^34^ In this investigation, the yield of a unit-based approach to identify individuals at risk of exposure was extremely low (0.2% yield), directly identifying only two new cases. Active surveillance is a recommended tool to perform case finding in epidemiologically significant organisms in healthcare settings.^35^ Defining more effective and efficient active surveillance strategies in outbreak and endemic settings is needed.

The outbreak plasmid we identified is of the *IncX3* replicon type, an emerging plasmid type frequently carrying *bla*_NDM_ genes.^36^ IncX5 NDM-5-encoding plasmids have been recovered from a multitude of bacterial hosts from clinical, animal, and environmental samples, highlighting their capability to exist in and move between a variety of settings.^37–40^ In addition to NDM-5, this plasmid also carries genes encoding a type IV secretion system, which likely enables efficient plasmid sharing between bacteria.

The presence of these plasmids poses a danger to hospitalized patients because they confer antimicrobial resistance and therefore complicate therapy, making their detection and control of the upmost importance. Although the outbreak plasmid was genetically most similar to plasmids isolated from China, the source of the plasmid and how it entered our hospital system is not known.

There are several limitations to this study. Furthermore, there was a one-year gap between Patient 1 and the other patients involved in the outbreak, during which the plasmid was not detected. suggesting the possibility that this patient was not the source of the outbreak. Only isolates requested by IP&C for WGS in reaction to a suspected outbreak, those chosen by the EDS-HAT algorithm for WGS surveillance, and those separately provided to the lab from the UPMC XDR Pathogens Lab underwent WGS. Therefore, it is likely that we have missed cases and consequently underestimated the true number of patients carrying NDM-producing organisms during the study period.

In conclusion, we characterized a complex outbreak involving multiple plasmid transfer and bacterial transmission events using traditional epidemiologic approaches and short- and long-read WGS. The implications of plasmid transfer events in healthcare settings on infection prevention and control are not yet fully understood. We currently run EDS-HAT in real time and typically detect at least one bacterial transmission outbreak each week using Illumina sequencing. EDS-HAT, however, was designed to detect bacterial transmission and not plasmid transfer. We are currently exploring the use of Illumina sequencing alone, in combination with a reference plasmid database, for inferring plasmid content. If successful, we plan to explore the incorporation of real-time detection of plasmid transfer events to determine how common these events are, with the ultimate goal of developing targeted interventions to halt the spread of multidrug-resistant plasmids and therefore improve patient safety.

### Contributors

LHH, GS, DVT, NJR designed the research study and oversaw the collection of data. MPG, EM, NJR, and DVT assisted with design, execution, and interpretation of bioinformatic/genomic analyses. AV, CB, AA, GS, NJR collected and interpreted IP&C and epidemiologic data. AJS, LP, KW, HC, VRS assisted with data collection, design, and interpretation of the analysis. All authors had full access to all the data in the study and NJR, AV, LHH, and GS verified the data. NJR and LHH wrote the initial draft of the manuscript, and all other authors reviewed subsequent drafts critically and approved the decision to submit the final draft for publication.

## Supporting information

Supplemental Figure 1: Representative Outbreak Plasmid Annotation

Supplemental Table 1: Plasmid Metadata

## DATA SHARING

Sequencing data for outbreak isolates and plasmids was made available on NCBI through BioProject PRJNA981541. Supplemental Table 1 lists accession information for all plasmid sequences analyzed in this study.

## Acknowledgements

We thank Ryan K. Shields for the provision of an important study isolate, and Vaughn Cooper for assistance with whole genome sequencing. This publication made use of the PubMLST website (https://pubmlst.org/) developed by Keith Jolley (Jolley & Maiden 2010, BMC Bioinformatics, 11:595) and sited at the University of Oxford. The development of that website was funded by the Wellcome Trust. Figure 1 was created with BioRender.com

## REFERENCES

1. San Millan A. Evolution of Plasmid-Mediated Antibiotic Resistance in the Clinical Context. Trends Microbiol. 2018 Dec;26(12):978–85.

2. Thomas CM, Nielsen KM. Mechanisms of, and barriers to, horizontal gene transfer between bacteria. Nat Rev Microbiol. 2005 Sep;3(9):711–21.

3. Snyder BM, Montague BT, Anandan S, Madabhushi AG, Pragasam AK, Verghese VP, et al. Risk factors and epidemiologic predictors of blood stream infections with New Delhi Metallo-b-lactamase (NDM-1) producing Enterobacteriaceae. Epidemiol Infect. 2019 Jan;147:e137.

4. Wu W, Feng Y, Tang G, Qiao F, McNally A, Zong Z. NDM Metallo-β-Lactamases and Their Bacterial Producers in Health Care Settings. Clin Microbiol Rev. 2019 Mar 20;32(2).

5. Sundermann AJ, Chen J, Kumar P, Ayres AM, Cho ST, Ezeonwuka C, et al. Whole-Genome Sequencing Surveillance and Machine Learning of the Electronic Health Record for Enhanced Healthcare Outbreak Detection. Clin Infect Dis. 2022 Aug 31;75(3):476– 82.

6. Sundermann AJ, Chen J, Miller JK, Martin EM, Snyder GM, Van Tyne D, et al. Whole-genome sequencing surveillance and machine learning for healthcare outbreak detection and investigation: A systematic review and summary. Antimicrob Steward Healthc Epidemiol. 2022 Jun 13;2(1):e91.

7. Shugart A, Mahon G, Epstein L, Huang JY, McAllister G, Lawsin A, et al. Changing US Epidemiology of NDM-Producing Carbapenem-Resistant Enterobacteriaceae, 2017– 2019. Infect Control Hosp Epidemiol. 2020 Oct;41(S1):s25–6.

8. Weber RE, Pietsch M, Frühauf A, Pfeifer Y, Martin M, Luft D, et al. IS26-Mediated Transfer of blaNDM-1 as the Main Route of Resistance Transmission During a Polyclonal, Multispecies Outbreak in a German Hospital. Front Microbiol. 2019 Dec 17;10:2817.

9. Lo S, Lolom I, Goldstein V, Petitjean M, Rondinaud E, Bunel-Gourdy V, et al. Simultaneous Hospital Outbreaks of New Delhi Metallo-β-Lactamase-Producing Enterobacterales Unraveled Using Whole-Genome Sequencing. Microbiol Spectr. 2022 Apr 27;10(2):e0228721.

10. Beukers AG, John MA, Davis R, Lee A, van Hal SJ. Hospital outbreak of New Delhi metallo-β-lactamase type-1 (NDM-1) in Salmonella enterica with inter-species plasmid transmission. J Hosp Infect. 2021 Nov;117:23–7.

11. Sundermann A, Griffith M, Srinivasa VR, Waggle K, Saul M, Ayres A, et al. Research and implementation of a whole-genome sequencing surveillance system for outbreak detection. ASHE. 2022 Jul;2(S1):s82–s82.

12. Mustapha MM, Srinivasa VR, Griffith MP, Cho S-T, Evans DR, Waggle K, et al. Genomic Diversity of Hospital-Acquired Infections Revealed through Prospective Whole-Genome Sequencing-Based Surveillance. mSystems. 2022 Jun 28;7(3):e0138421.

13. CLSI. Performance Standards for Antimicrobial Susceptibility Testing. 33rd ed. CLSI supplement M100. clinical and Laboratory Standards Institute; 2023.

14. Jain C, Rodriguez-R LM, Phillippy AM, Konstantinidis KT, Aluru S. High throughput ANI analysis of 90K prokaryotic genomes reveals clear species boundaries. Nat Commun. 2018 Nov 30;9(1):5114.

15. Bankevich A, Nurk S, Antipov D, Gurevich AA, Dvorkin M, Kulikov AS, et al. SPAdes: a new genome assembly algorithm and its applications to single-cell sequencing. J Comput Biol. 2012 May;19(5):455–77.

16. Seemann T. Prokka: rapid prokaryotic genome annotation. Bioinformatics. 2014 Jul 15;30(14):2068–9.

17. Mikheenko A, Prjibelski A, Saveliev V, Antipov D, Gurevich A. Versatile genome assembly evaluation with QUAST-LG. Bioinformatics. 2018 Jul 1;34(13):i142–50.

18. Wick RR, Judd LM, Gorrie CL, Holt KE. Unicycler: Resolving bacterial genome assemblies from short and long sequencing reads. PLoS Comput Biol. 2017 Jun 8;13(6):e1005595.

19. Carattoli A, Zankari E, García-Fernández A, Voldby Larsen M, Lund O, Villa L, et al. In silico detection and typing of plasmids using PlasmidFinder and plasmid multilocus sequence typing. Antimicrob Agents Chemother. 2014 Jul;58(7):3895–903.

20. Page AJ, Cummins CA, Hunt M, Wong VK, Reuter S, Holden MTG, et al. Roary: rapid large-scale prokaryote pan genome analysis. Bioinformatics. 2015 Nov 15;31(22):3691– 3.

21. Stamatakis A. RAxML version 8: a tool for phylogenetic analysis and post-analysis of large phylogenies. Bioinformatics. 2014 May 1;30(9):1312–3.

22. Evans D, Sundermann A, Griffith M, Rangachar Srinivasa V, Mustapha M, Chen J, et al. Empirically derived sequence similarity thresholds to study the genomic epidemiology of plasmids shared among healthcare-associated bacterial pathogens. EBioMedicine. 2023 Jun 29;93:104681.

23. Sundermann AJ, Chen J, Miller JK, Saul MI, Shutt KA, Griffith MP, et al. Outbreak of Pseudomonas aeruginosa Infections from a Contaminated Gastroscope Detected by Whole Genome Sequencing Surveillance. Clin Infect Dis. 2021 Aug 2;73(3):e638–42.

24. Sundermann AJ, Babiker A, Marsh JW, Shutt KA, Mustapha MM, Pasculle AW, et al. Outbreak of Vancomycin-resistant Enterococcus faecium in Interventional Radiology: Detection Through Whole-genome Sequencing-based Surveillance. Clin Infect Dis. 2020 May 23;70(11):2336–43.

25. Ward DV, Hoss AG, Kolde R, van Aggelen HC, Loving J, Smith SA, et al. Integration of genomic and clinical data augments surveillance of healthcare-acquired infections. Infect Control Hosp Epidemiol. 2019 Jun;40(6):649–55.

26. Berbel Caban A, Pak TR, Obla A, Dupper AC, Chacko KI, Fox L, et al. PathoSPOT genomic epidemiology reveals under-the-radar nosocomial outbreaks. Genome Med. 2020 Nov 16;12(1):96.

27. Jakharia KK, Ilaiwy G, Moose SS, Waga M, Appalla L, McAlduff JD, et al. Use of whole-genome sequencing to guide a Clostridioides difficile diagnostic stewardship program. Infect Control Hosp Epidemiol. 2019 Jul;40(7):804–6.

28. Gona F, Comandatore F, Battaglia S, Piazza A, Trovato A, Lorenzin G, et al. Comparison of core-genome MLST, coreSNP and PFGE methods for Klebsiella pneumoniae cluster analysis. Microb Genom. 2020 Apr;6(4).

29. Rose R, Nolan DJ, Moot S, Rodriguez C, Cross S, McCarter YS, et al. Molecular surveillance of methicillin-resistant Staphylococcus aureus genomes in hospital unexpectedly reveals discordance between temporal and genetic clustering. Am J Infect Control. 2021 Jan;49(1):59–64.

30. Marmor A, Daveson K, Harley D, Coatsworth N, Kennedy K. Two carbapenemase-producing Enterobacteriaceae outbreaks detected retrospectively by whole-genome sequencing at an Australian tertiary hospital. Infection, Disease & Health. 2020 Feb;25(1):30–3.

31. Sherry NL, Lane CR, Kwong JC, Schultz M, Sait M, Stevens K, et al. Genomics for Molecular Epidemiology and Detecting Transmission of Carbapenemase-Producing Enterobacterales in Victoria, Australia, 2012 to 2016. J Clin Microbiol. 2019 Sep;57(9).

32. Raven KE, Gouliouris T, Brodrick H, Coll F, Brown NM, Reynolds R, et al. Complex Routes of Nosocomial Vancomycin-Resistant Enterococcus faecium Transmission Revealed by Genome Sequencing. Clin Infect Dis. 2017 Apr 1;64(7):886–93.

33. Kwong JC, Lane CR, Romanes F, Gonçalves da Silva A, Easton M, Cronin K, et al. Translating genomics into practice for real-time surveillance and response to carbapenemase-producing Enterobacteriaceae: evidence from a complex multi-institutional KPC outbreak. PeerJ. 2018 Jan 3;6:e4210.

34. van Loon K, Voor In ’t Holt AF, Vos MC. A Systematic Review and Meta-analyses of the Clinical Epidemiology of Carbapenem-Resistant Enterobacteriaceae. Antimicrob Agents Chemother. 2018 Jan;62(1).

35. National Center for Emerging and Zoonotic Infectious Diseases (U.S.), Division of Healthcare Quality Promotion. “Guidance for control of Carbapenem-resistant Enterobacteriaceae (CRE); 2012 CRE toolkit”, 2012 [Internet]. [cited 2023 Jul 5]. Available from: https://stacks.cdc.gov/view/cdc/13205

36. Ma T, Fu J, Xie N, Ma S, Lei L, Zhai W, et al. Fitness Cost of blaNDM-5-Carrying p3R-IncX3 Plasmids in Wild-Type NDM-Free Enterobacteriaceae. Microorganisms. 2020 Mar 7;8(3).

37. Baomo L, Lili S, Moran RA, van Schaik W, Chao Z. Temperature-Regulated IncX3 Plasmid Characteristics and the Role of Plasmid-Encoded H-NS in Thermoregulation. Front Microbiol. 2021;12:765492.

38. Zhou H, Zhang K, Chen W, Chen J, Zheng J, Liu C, et al. Epidemiological characteristics of carbapenem-resistant Enterobacteriaceae collected from 17 hospitals in Nanjing district of China. Antimicrob Resist Infect Control. 2020 Jan 13;9(1):15.

39. Nigg A, Brilhante M, Dazio V, Clément M, Collaud A, Gobeli Brawand S, et al. Shedding of OXA-181 carbapenemase-producing Escherichia coli from companion animals after hospitalisation in Switzerland: an outbreak in 2018. Euro Surveill. 2019 Sep;24(39).

40. Zhai R, Fu B, Shi X, Sun C, Liu Z, Wang S, et al. Contaminated in-house environment contributes to the persistence and transmission of NDM-producing bacteria in a Chinese poultry farm. Environ Int. 2020 Jun;139:105715.

