## Supplemental Figure 1: Representative Outbreak Plasmid Annotation for "Genomic Epidemiologic Investigation of a Multispecies Hospital Outbreak of NDM-5-Producing Enterobacterales Infections"

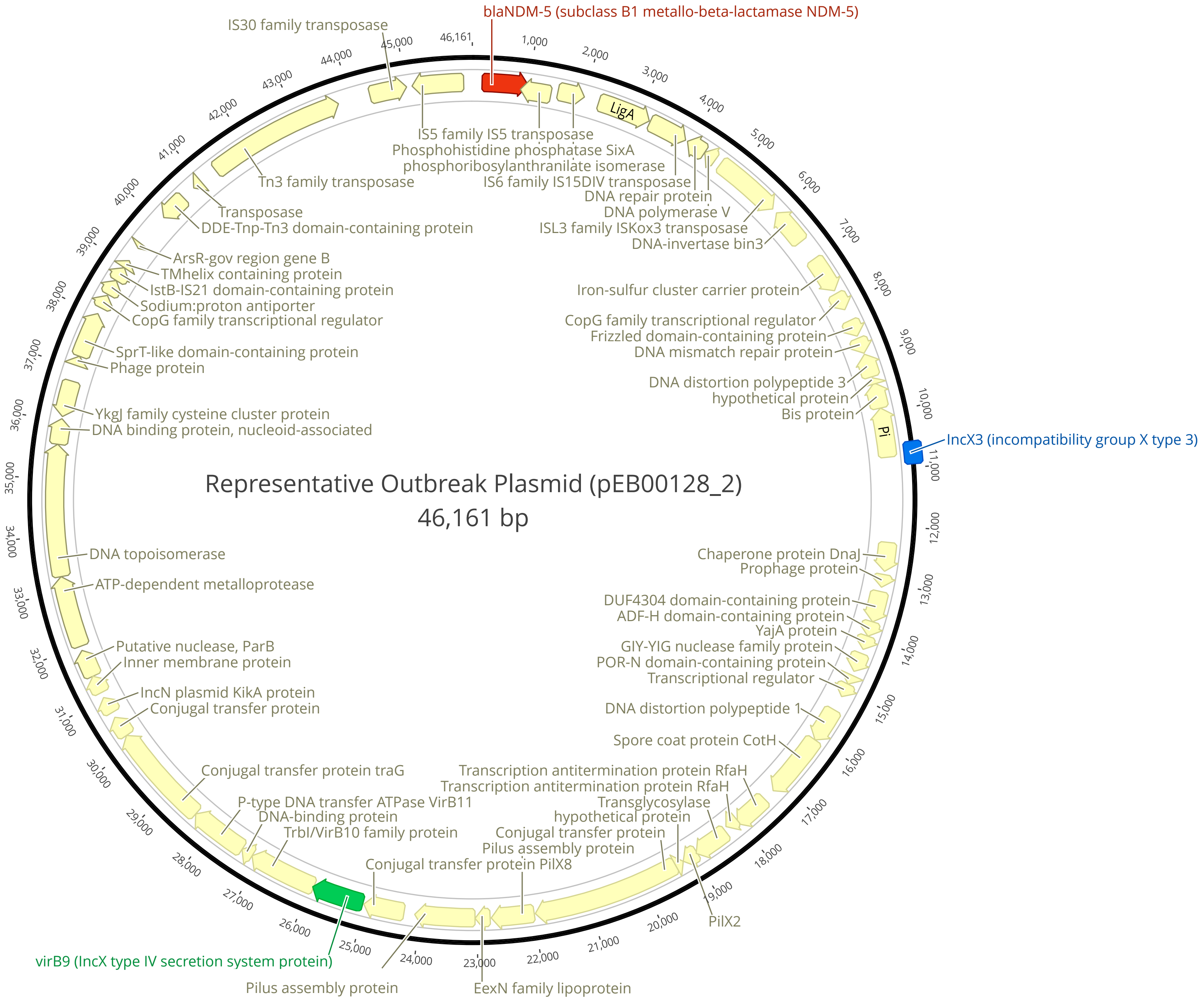


**Supplemental Figure 1. Representative outbreak plasmid annotation.** The predominant form of the *IncX3 bla*$_{\text{NDM-5}}$-encoding outbreak plasmid sequence is shown. Annotations were created using Bakta and visualized in Geneious. Important genes are highlighted; *bla*$_{\text{NDM-5}}$ (subclass B1 metallo-beta-lactamase NDM-5) is shown in red, *IncX3* (incompatibility group X type 3) is shown in blue, and *virB9* (*IncX* type IV secretion system protein) is shown in green.
