## Supplemental Table 1: Plasmid Metadata for "Genomic Epidemiologic Investigation of a Multispecies Hospital Outbreak of NDM-5-Producing Enterobacterales Infections"

| Plasmid ID | NCBI Accession No. | Similarity ScoreA | Sequence Length (bp) | Publication SourceB | Species | ML STC | Rep Gene | T4SS D | NDM Type | Isolation Source | Country of Isolation | Year |
|---|---|---|---|---|---|---|---|---|---|---|---|---|
| **EB00131** | SAMN35677778 | 100.00 | 46161 | This Study | *E. hormaechei* | 45 | IncX3 | Yes | blaNDM-5 | Human | United States | 2022 |
| **EB00132** | SAMN35677779 | 100.00 | 46161 | This Study | *E. hormaechei* | 45 | IncX3 | Yes | blaNDM-5 | Human | United States | 2022 |
| **MULT0069** | SAMN35677780 | 100.00 | 46161 | This Study | *M. morganii* | NA | IncX3 | Yes | blaNDM-5 | Human | United States | 2022 |
| **PR00728** | SAMN35677781 | 100.00 | 46161 | This Study | *P. mirabilis* | NA | IncX3 | Yes | blaNDM-5 | Human | United States | 2022 |
| **EB00148** | SAMN35677782 | 100.00 | 46161 | This Study | *E. hormaechei* | 45 | IncX3 | Yes | blaNDM-5 | Human | United States | 2022 |
| **SURV0398** | SAMN35677783 | 100.00 | 46161 | This Study | *E. coli* | 131 | IncX3 | Yes | blaNDM-5 | Human | United States | 2022 |
| **EB00150** | SAMN35677784 | 100.00 | 46161 | This Study | *K. aerogenes* | 239 | IncX3 | Yes | blaNDM-5 | Human | United States | 2022 |
| **EB00147** | SAMN35677785 | 100.00 | 46161 | This Study | *E. hormaechei* | 45 | IncX3 | Yes | blaNDM-5 | Human | United States | 2022 |
| **EB00128** | SAMN35677786 | 100.00 | 46161 | This Study | *K. aerogenes* | NA | IncX3 | Yes | blaNDM-5 | Human | United States | 2022 |
| **ENTC0964** | SAMN35677787 | 73.62 | 45198 | This Study | *E. hormaechei* | NA | IncX3 | Yes | blaNDM-5 | Human | United States | 2021 |
| **EB00158** | SAMN35677788 | 100.00 | 46161 | This Study | *E. hormaechei* | 45 | IncX3 | Yes | blaNDM-5 | Human | United States | 2022 |
| **EB00153** | SAMN35677789 | 100.00 | 46161 | This Study | *K. aerogenes* | 137 SLV | IncX3 | Yes | blaNDM-5 | Human | United States | 2022 |
| **EB00162** | SAMN35677790 | 59.39 | 55781 | This Study | *E. hormaechei* | 45 | IncX3 | Yes | blaNDM-5 | Human | United States | 2022 |
| **EB00163** | SAMN35677791 | 100.00 | 46161 | This Study | *E. hormaechei* | 145 | IncX3 | Yes | blaNDM-5 | Human | United States | 2022 |
| **KLP00703** | SAMN35677792 | 100.00 | 46161 | This Study | *R. ornithinolytica* | NA | IncX3 | Yes | blaNDM-5 | Human | United States | 2022 |
| **EB00175** | SAMN35677793 | 100.00 | 46161 | This Study | *E. roggenkampii* | 997 | IncX3 | Yes | blaNDM-5 | Human | United States | 2023 |
| **EB00176** | SAMN35677794 | 100.00 | 46161 | This Study | *E. roggenkampii* | 997 | IncX3 | Yes | blaNDM-5 | Human | United States | 2023 |
| **EB00178** | SAMN35677795 | 100.00 | 46161 | This Study | *E. roggenkampii* | 997 | IncX3 | Yes | blaNDM-5 | Human | United States | 2023 |
| **EC01307** | SAMN35677796 | 100.00 | 46161 | This Study | *E. coli* | 162 | IncX3 | Yes | blaNDM-5 | Human | United States | 2023 |
| **pSCK27-NDM5** | MT663954.1 | 100.00 | 46161 | Zhu W., et al. 2020 mSphere. doi: 10.1128/mSphere.00917-20 | *K. pneumoniae* | 48 | IncX3 | Yes | blaNDM-5 | Human | China | 2016-2018 |
| **pABC233-NDM-5** | MK372390.1 | 100.00 | 46161 | Mouftah SF., et al. 2019 Infect Drug Resist. doi: 10.2147/IDR.S210554 | *E. coli* | 167 | IncX3 | Yes | blaNDM-5 | Human | United Arab Emirates | 2013 |
| **pABC384-NDM-5** | MK372389.1 | 100.00 | 46161 | Mouftah SF., et al. 2019 Infect Drug Resist. doi: 10.2147/IDR.S210554 | *E. coli* | 1284 | IncX3 | Yes | blaNDM-5 | Human | United Arab Emirates | 2014 |
| **pNDM-HK2998** | MH234508.1 | 100.00 | 46161 | Wang Y., et al. 2018 Front. Microbiol. doi: 10.3389/fmicb.2018.02272 | *K. pneumoniae* | NA | IncX3 | Yes | blaNDM-5 | Human | China | 2016 |
| **pNDM-HK3218** | MH234507.1 | 100.00 | 46161 | Wang Y., et al. 2018 Front. Microbiol. doi: 10.3389/fmicb.2018.02272 | *E. coli* | NA | IncX3 | Yes | blaNDM-5 | Human | China | 2016 |

| | | | | | | | | | | | | |
|---|---|---|---|---|---|---|---|---|---|---|---|---|
| **pNDM-HK3473** | MH234506.1 | 100.00 | 46161 | Wang Y., et al. 2018 Front. Microbiol. doi: 10.3389/fmicb.2018.02272 | *E. cloacae* | NA | IncX3 | Yes | blaNDM-5 | Human | China | 2016 |
| **pNDM-HK3706** | MH234504.1 | 99.90 | 46161 | Wang Y., et al. 2018 Front. Microbiol. doi: 10.3389/fmicb.2018.02272 | *K. pneumoniae* | NA | IncX3 | Yes | blaNDM-5 | Human | China | 2016 |
| **pNDM-HK3712** | MH234503.1 | 99.99 | 46161 | Wang Y., et al. 2018 Front. Microbiol. doi: 10.3389/fmicb.2018.02272 | *E. coli* | NA | IncX3 | Yes | blaNDM-5 | Human | China | 2016 |
| **pNDM-HK3816** | MH234501.1 | 99.92 | 46161 | Wang Y., et al. 2018 Front. Microbiol. doi: 10.3389/fmicb.2018.02272 | *K. pneumoniae* | NA | IncX3 | Yes | blaNDM-5 | Human | China | 2016 |
| **pNDM-HK3819** | MH234500.1 | 100.00 | 46161 | Wang Y., et al. 2018 Front. Microbiol. doi: 10.3389/fmicb.2018.02272 | *K. pneumoniae* | NA | IncX3 | Yes | blaNDM-5 | Human | China | 2016 |
| **pNDM-HK3855** | MH234498.1 | 100.00 | 46161 | Wang Y., et al. 2018 Front. Microbiol. doi: 10.3389/fmicb.2018.02272 | *E. coli* | NA | IncX3 | Yes | blaNDM-5 | Human | China | 2016 |
| **pNDM-HK3871** | MH234497.1 | 100.00 | 46161 | Wang Y., et al. 2018 Front. Microbiol. doi: 10.3389/fmicb.2018.02272 | *E. coli* | NA | IncX3 | Yes | blaNDM-5 | Human | China | 2016 |
| **pEC463-NDM5** | MG545911.1 | 99.96 | 46145 | Xi L., et al. 2018 Antimicrob Resist Infect Control. doi: 10.1186/s13756-018-0349-6 | *E. coli* | 361 | IncX3 | Yes | blaNDM-5 | Human | China | 2016 |
| **pP768-NDM5** | MF547510.1 | 100.00 | 46161 | Ho PL., et al. 2018 Antimicrb Agents Chemother. doi: 10.1128/AAC.02295-17 | *E. coli* | 117 | IncX3 | Yes | blaNDM-5 | Pig | China | 2016 |
| **pP785-NDM5** | MF547509.1 | 100.00 | 46161 | Ho PL., et al. 2018 Antimicrb Agents Chemother. doi: 10.1128/AAC.02295-17 | *E. coli* | 7512 | IncX3 | Yes | blaNDM-5 | Pig | China | 2016 |
| **pP855-NDM5** | MF547508.1 | 100.00 | 46161 | Ho PL., et al. 2018 Antimicrb Agents Chemother. doi: 10.1128/AAC.02295-17 | *E. coli* | 7511 | IncX3 | Yes | blaNDM-5 | Pig | China | 2017 |
| **pP788A-NDM5** | MF547507.1 | 100.00 | 46161 | Ho PL., et al. 2018 Antimicrb Agents Chemother. doi: 10.1128/AAC.02295-17 | *E. coli* | 1286 | IncX3 | Yes | blaNDM-5 | Pig | China | 2016 |
| **pECNDM101** | KX507346.1 | 99.97 | 46165 | Kong LH., et al. 2017 Antimirob Agents Chemother. doi :10.1128/AAC.02167-16 | *E. coli* | NA | IncX3 | Yes | blaNDM-5 | Pig | China | 2015 |
| **pNDM5_IncX3** | KU761328.1 | 100.00 | 46161 | Li, A., et al. 2016 Antimicrob Agents Chemother. doi: 10.1128/AAC.00550-16 | *K. pneumoniae* | 25 | IncX3 | Yes | blaNDM-5 | Human | China | 2015 |
| **pNDM-QD29** | KU167609.1 | 99.99 | 46161 | Zhu, Y-Q., et al. 2016 Sci Rep. doi: 10.1038/srep29934 | *E. coli* | 167 | IncX3 | Yes | blaNDM-5 | Human | China | 2013 |
| **pNDM-QD28** | KU167608.1 | 100.00 | 46161 | Zhu, Y-Q., et al. 2016 Sci Rep. doi: 10.1038/srep29934 | *E. coli* | 167 | IncX3 | Yes | blaNDM-5 | Human | China | 2013 |
| **pEc1929** | KT824791.1 | 99.99 | 46164 | Chen, D., et al. 2016 J Antimicrob Chemother. doi: 10.1093/jac/dkv352 | *E. coli* | NA | IncX3 | Yes | blaNDM-5 | Human | China | 2014 |
| **pSCZP2** | CP051221.1 | 99.99 | 46160 | He, J., et al. 2020 Front Microbiol. doi: 10.3389/fmicb.2020.02049 | *E. coli* | 533 | IncX3 | Yes | blaNDM-5 | Pig | China | 2019 |
| **p3R-4** | CP049352.1 | 99.86 | 46118 | Ma, T., et al. 2020 Microorganisms. doi: 10.3390/microorganisms8030377 | *E. coli* | 156 | IncX3 | Yes | blaNDM | Poultry | China | 2015 |
| **pK19CRE38** | CP049051.1 | 100.00 | 46161 | Moon, D.C., et al. Source Unknown | *E. coli* | 1721 | IncX3 | Yes | blaNDM-5 | Dog | South Korea | 2019 |
| **pNDM5-GZEC065** | CP048028.1 | 99.98 | 46161 | Lin, Y., et al. 2020 Infect Genet Evol. doi: 10.1016/j.meegid.2020.104499 | *E. coli* | 156 | IncX3 | Yes | blaNDM-5 | Human | China | 2017 |
| **pNDM5_025970** | CP036179.1 | 99.99 | 46161 | Long, H., et al. Source Unknown. | *E. coli* | 167 | IncX3 | Yes | blaNDM-5 | Human | China | 2017 |
| **pNDM5_020032** | CP034965.1 | 99.99 | 46161 | Liu, Z. et al. Source Unknown. | *E. coli* | 410 | IncX3 | Yes | blaNDM-5 | Human | China | 2016 |
| **pNDM5_020026** | CP034957.1 | 100.00 | 46161 | Feng, Y., et al. 2019 Communications Biology. doi: 10.1038/s42003-019-0569-1 | *E. coli* | 410 | IncX3 | Yes | blaNDM-5 | Human | China | 2016 |

| | | | | | | | | | | | | |
|---|---|---|---|---|---|---|---|---|---|---|---|---|
| **pL100-4** | CP034748.1 | 99.99 | 46161 | Liu, Z., et al. 2019 Front. Microbiol. Doi: 10.3389/fmicb.2019.02002 | *E. coli* | 48 | IncX3 | Yes | blaNDM-5 | Goose | China | 2018 |
| **pL65-9** | CP034744.1 | 100.00 | 46161 | Liu, Z., et al. 2019 Front. Microbiol. Doi: 10.3389/fmicb.2019.02002 | *E. coli* | 3076 | IncX3 | Yes | blaNDM-5 | Goose | China | 2018 |
| **pL41-1-4** | CP034730.1 | 99.93 | 46238 | Liu, Z., et al. 2019 Front. Microbiol. Doi: 10.3389/fmicb.2019.02002 | *E. coli* | 48 | IncX3 | Yes | blaNDM-5 | Goose | China | 2018 |
| **pNDM5_020031** | CP033399.1 | 100.00 | 46161 | Liu, L., et al. Source Unknown. | *E. coli* | 410 | IncX3 | Yes | blaNDM-5 | Human | China | 2016 |
| **pNDM5_020022** | CP032889.1 | 100.00 | 46161 | Liu, L., et al. Source Unknown. | *E. coli* | 156 | IncX3 | Yes | blaNDM-5 | Human | China | 2016 |
| **pNDM5_020001** | CP032424.1 | 100.00 | 46161 | Liu, L., et al. Source Unknown. | *E. coli* | 410 | IncX3 | Yes | blaNDM-5 | Human | China | 2016 |
| **pTB203** | CP029245.1 | 99.99 | 46161 | Tang, B., et. 2019 BMC Microbiology. doi: 10.1186/s12866-019-1454-2 | *E. coli* | 156 | IncX3 | Yes | blaNDM-5 | Poultry | China | 2017 |
| **pNDM5_005784** | CP028577.1 | 100.00 | 46161 | Feng, Y., et al. Source Unknown. | *E. coli* | 617 | IncX3 | Yes | blaNDM-5 | Human | China | 2014 |
| **pNDM5_005237** | CP026577.2 | 100.00 | 46161 | Feng, Y., et al. Source Unknown. | *E. coli* | 167 | IncX3 | Yes | blaNDM-5 | Human | China | 2014 |
| **pCREC-591_4** | CP024825.1 | 99.99 | 46161 | Yoon, EJ.,et al. 2018 Front Microbiol. doi: 10.3389/fmicb.2018.00571 | *E. coli* | 101 | IncX3 | Yes | blaNDM-5 | Human | South Korea | 2015 |
| **pK518_NDM5** | CP023188.1 | 99.97 | 46146 | Zheng B., et al. Source Unknown. | *K. michiganensis* | NA | IncX3 | Yes | blaNDM-5 | Human | China | 2017 |
| **pK516_NDM5** | CP022351.1 | 99.96 | 46165 | Zheng B., et al. Source Unknown. | *K. michiganensis* | NA | IncX3 | Yes | blaNDM-5 | Human | China | 2017 |
| **p165** | CP020512.1 | 99.99 | 46159 | Hoffman, M., et al. Source Unknown. | *E. coli* | 101 | IncX3 | Yes | blaNDM-5 | Human | United States | 2015 |
| **p1493-2** | CP019073.1 | 99.96 | 46161 | Ho, P.L., et al. Source Unknown. | *E. coli* | 167 | IncX3 | Yes | blaNDM-5 | Human | China | 2013 |
| **NUHL24835** | CP014006.1 | 100.00 | 46161 | Mei, YF., et al. 2017 Front. Microbiol. doi: 10.3389/fmicb.2017.00335 | *K. pneumoniae* | 14 | IncX3 | Yes | blaNDM-5 | Human | China | 2014 |
| **pMTY18781-5** | AP023210.1 | 100.00 | 46161 | Ito, Y., et al. 2022 Microb Drug Resist. Doi: 10.1089/mdr.2021.0197 | *E. coli* | 2040 | IncX3 | Yes | blaNDM-5 | Human | Japan | 2018 |
| **pWP8-S18-CRE-02_4** | AP022249.1 | 99.99 | 46158 | Sekizuka, T., et al. 2022 Antibiotics (Basel). doi: 10.3390/antibiotics11101283 | *E. coli* | 542 | IncX3 | Yes | blaNDM-5 | Wastewater treatment plant-environment | Japan | 2018 |
| **pGSH8M-2-4** | AP019679.1 | 99.99 | 44811 | Sekizuka, T., et al. 2019 Infect Drug Resist. doi: 10.2147/IDR.S215273 | *E. coli* | 542 | IncX3 | Yes | blaNDM-5 | wastewater treatment plant-environment | Japan | 2018 |

[A] Similarity Score was caculated based on the percent identity multipled by percent coverage to reference study plasmid PR00728.
[B] Plasmids from unknown sources are noted as such with the NCBI submitter's name.
[C] Isolates without an available mutilocus sequence type (MLST) scheme or lack of publically available information on MLST are denoted as not available (NA).
[D] Plasmids contain machinery for a Type IV Secretion System.
